# CHARACTERISTICS AND OUTCOMES OF INDIVIDUALS WITH COVID-19: EVIDENCE FROM A HOSPITAL BASED STUDY

**DOI:** 10.1101/2022.05.13.22274812

**Authors:** P. Simushi, M. Zambwe, P.J. Chipimo

**Affiliations:** School of Medicine and Health Sciences, University of Lusaka, Lusaka, Zambia; Benefits Department, Workers Compensation Fund Control Board, Lusaka, Zambia; Surveillance and Disease Intelligence Cluster, Zambia National Public Health Institute, Lusaka, Zambia

**Author notes:** **<u>CORRESPONDING AUTHOR DETAILS</u> Name:** Mowa Zambwe, **Address:** Workers Compensation Fund Control Board, Lusaka, Zambia, **Phone Number:** **, **Email ID:**. **Guarantor of Submission:** The corresponding author is the guarantor of submission.

## Abstract

**Objective:** To determine the characteristics and outcomes of Covid-19 patients at Livingstone teaching hospital.

**Methods:** A database cross sectional review of COVID 19 patients at Livingstone Teaching Hospital. Data on demographics and clinical characteristics were obtained along with the comorbidities presented with by the patients using a structured data collection form. Data were analysed using STATA 64. Mann-Whitney and t-test were used on continuous independent variables. Chi-square test was used to determine associations between two categorical variables. Logistic regression was used to control for confounders.

**Results:** A total of 222 (62.54%) were male and 133 (37.46%) were female. Discharged were 274 (77.18%), while 81 (22.82%) died. Among those who died were the older clients with a median age of 65 (p <0.001). The median interquartile range (IQR)] age was 48.5 years. Patients presented with a cough 180 (50.7%), chest pain 123 (34.65%) and shortness of breath 121 (34.04%). Statistically significant comorbidities recorded included Hypertension 121 (34.08%), Diabetes mellitus 69 (19.44%), and HIV 38 (10.7%). The most prevalent underlying condition observed was hypertension 121 (34.08).

**Conclusion:** HIV positive and diabetics had an increased odds of succumbing to COVID-19 death. It is recommended that targeted policies should be considered the risky groups.

## Introduction

The Severe acute respiratory syndrome coronavirus 2 (Sars-CoV-2) responsible for causing the COVID-19 infection is a novel Coronavirus. It is of zoonotic origins and belongs to the betacoronavirus genra with similar phylogenetic characteristics as the SARS-CoV [1]. It was described in December, 2019 by the World Health Organisation (WHO) as an acute respiratory disease infection responsible for a cluster of atypical pneumonia cases reported two weeks earlier in Wuhan city, Hubei province, in China [1–3].

WHO indicated the COVID-19 pandemic to be of public health emergency and of international concern that needed extensive resources directed at containing and controlling the spread of the virus as a surge in the number of cases was observed [1,2]. The severity of some of the cases of Covid-19 infection mimicked that of SARS-CoV, however, it was observed that the SARS-CoV-2 virus is not similar to other coronaviruses that usually spread in human beings responsible for causing common cold to severe respiratory distress syndrome [4,5]. The SARS-CoV-2 has shown to overwhemingly surpass SARS and MERS in terms of the number of people infected and the spatial range of epidemic areas [4].

By the second week of June the number of COVID-19 cases surpassed 200,000 and had escalated to 400,000 by the 6^th^ of July,2020 [6]. On the 14^th^ of February, 2020 and 16^th^ of March, 2020 the first case of Sars-cov-2 infection was reported in Egypt and Zambia respectively [7,8]. As opposed to the prediction by public health experts on COVID-19 in Africa, worst outcomes, the incidences, hospitalization and mortality rates recorded have been to a minimal as compared to other continents [6].

Human population at the time of the pandemic had no direct immunological experience with Sars-CoV-2, leaving the population vulnerable to infection and disease hence the devastating outcomes experienced globally [9–11]. This led to a surge in cases recorded, hospitalization and an elevation in mortality across the globe thus the desired need to determine the characteristics and outcomes of Covid-19 clients tailored to our Zambian settings.

## Methods

This was a cross sectional study based on analysis of secondary data. The data was collected using a structured data collection form. Medical records and data for the admitted clients with laboratory confirmed Covid-19 PCR and/or RDT positive results recorded from January 2020 to September 2021 were collected. The study excluded clients’ files without a COVID-19 positive PCR result or COVID-19 RDT result. It excluded all those that were not admitted at the Livingstone Teaching Hospital COVID-19 isolation ward.

We conducted a census of all patients admitted at Livingstone Teaching Hospital after permission had been granted by the University of Lusaka, School of Medicine & Health Sciences Research Ethics Committee, the National Health Research Authority (NHRA) and Mulungushi University School of Medicine and Health Sciences research ethics committee (MUSoMHS-REC).

Data was analysed using STATA 64 to produce descriptive statistics. Mann-Whitney and t-test were used on continuous independent variables between those with and those without outcome. Chi-square test was used to determine associations between two categorical variables. Logistic regression was used to control for confounding and determine the contribution of each variable towards the outcome. Sub analysis of the outcome variable was conducted to determine the relationship between each underlying condition and the predicting variables.

## Results

### Basic characteristics of study population

**Table I:** Demographics and clinical characteristics

| Characteristics | COVID-19 Participants |  |
| --- | --- | --- |
|  | n | % |
| *Demographics* |  |  |
| Age, Q1MQ2 | 340 | 37 48.5 65 |
| Gender, n (%) |  |  |
| Male | 222 | 62.54 |
| Female | 133 | 37.46 |
| *Clinical characteristics* |  |  |
| Fever, n (%) |  |  |
| No | 217 | 61.13 |
| Yes | 138 | 38.87 |
| Chest pain, n (%) |  |  |
| No | 232 | 65.35 |
| Yes | 123 | 34.65 |
| Cough, n (%) |  |  |
| No | 175 | 49.3 |
| Yes | 180 | 50.7 |
| Shortness of breath, n (%) |  |  |
| No | 234 | 65.92 |
| Yes | 121 | 34.08 |

Among the study population, 222 (62.54%) were male and 133 (37.46%) were female. The median age, interquartile range (IQR)] age was 48.5. Upon admission clinical presentation were recorded for all clients that were confirmed positive with the Sars-Cov-2. Significant characteristic observed on admission included cough 180 (50.7%), followed by fever 138 (38.87%) then chest pain with 123 (34.65%). From the study population of 355 clients, significant comorbidities; HIV positive clients recorded were 38 (10.7%), hypertension 121 (34.08), Diabetes mellitus clients were 69 (19.44%).

### Underlying conditions in COVID-19 Participants

**Table II:** Descriptive statistics of underlying conditions in COVID-19 Participants

| **Conditions** | **Frequency**<br>**n 355** | **Percentage**<br>**%** |
| --- | --- | --- |
| **HIV** |  |  |
| Positive | 38 | 10.7 |
| **DM, n (%)** |  |  |
| Yes | 69 | 19.44 |
| **HTN, n (%)** |  |  |
| Yes | 121 | 34.08 |
| **Outcome, n (%)** |  |  |
| Discharged | 274 | 77.18 |
| Death | 81 | 22.82 |

### Bivariate relationship between the outcome and demographics, clinical characteristics and underlying condition

Table III shows the relationship between outcome and demographics, clinical characteristics and underlying conditions. Majority of the Clients that died due to COVID-19 were significantly older than clients that survived (median age; 65 vs 45; p <0.001). The proportion of clients that died was higher among the male as compared to females [58 (71.60) vs 23(28.49]. There was a statistical difference in clients that presented with a clinical characteristic of shortness of breath between those that died 44.44% and those that survived 31.02% with a p-value of 0.025. The percentage of clients admitted with a known HIV positive status that died (18.52%) was statistically higher than those that survived (8.23%), p-value 0.013. Individuals with Diabetes Mellitus that died (33.33%) compared to those that survived (15.33%) was significant, p-value < 0.001 and clients who were hypertensive that died (50.62%) compared to those that survived (29.20%) was high, p-value <0.001.

**Table III:**
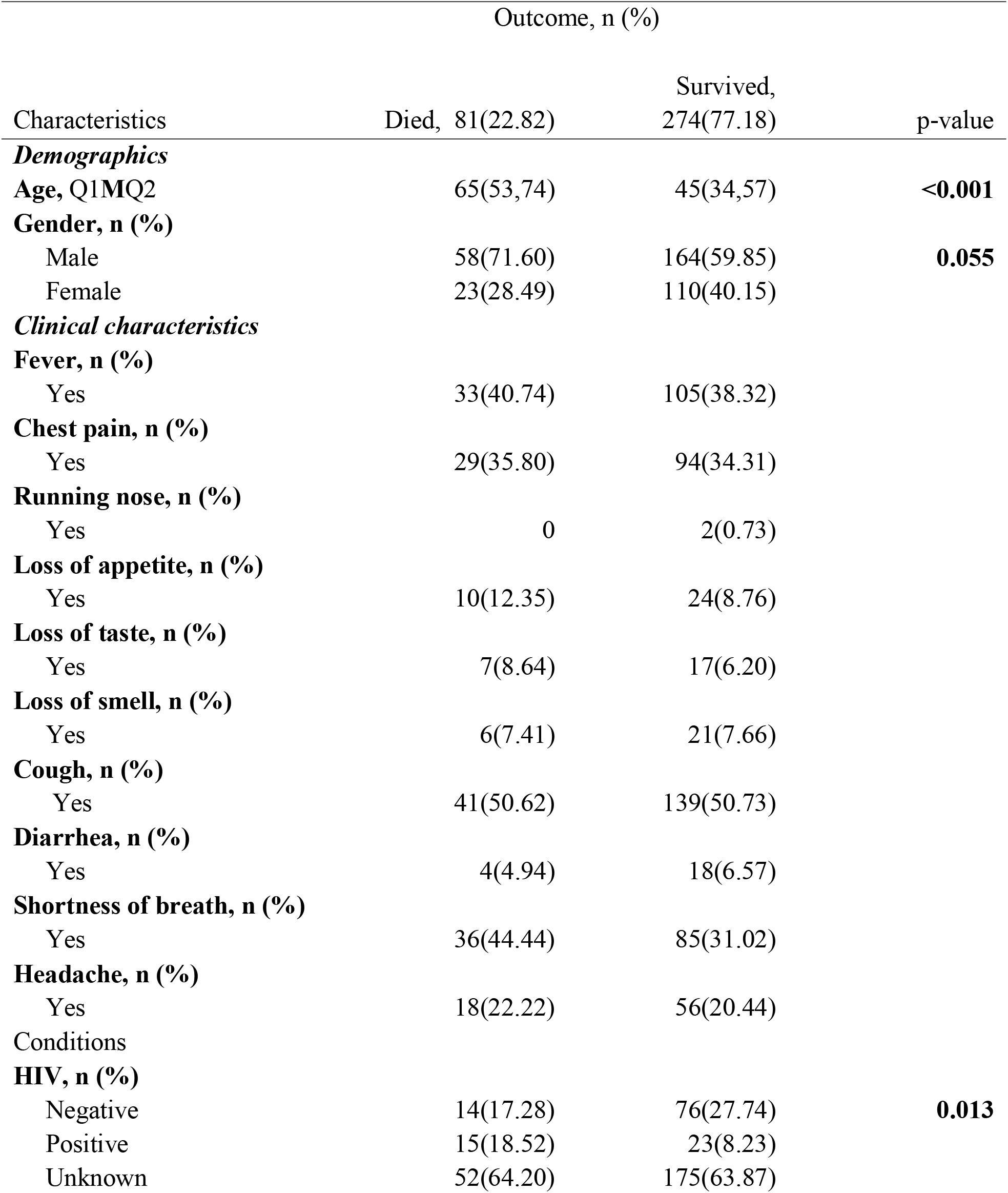
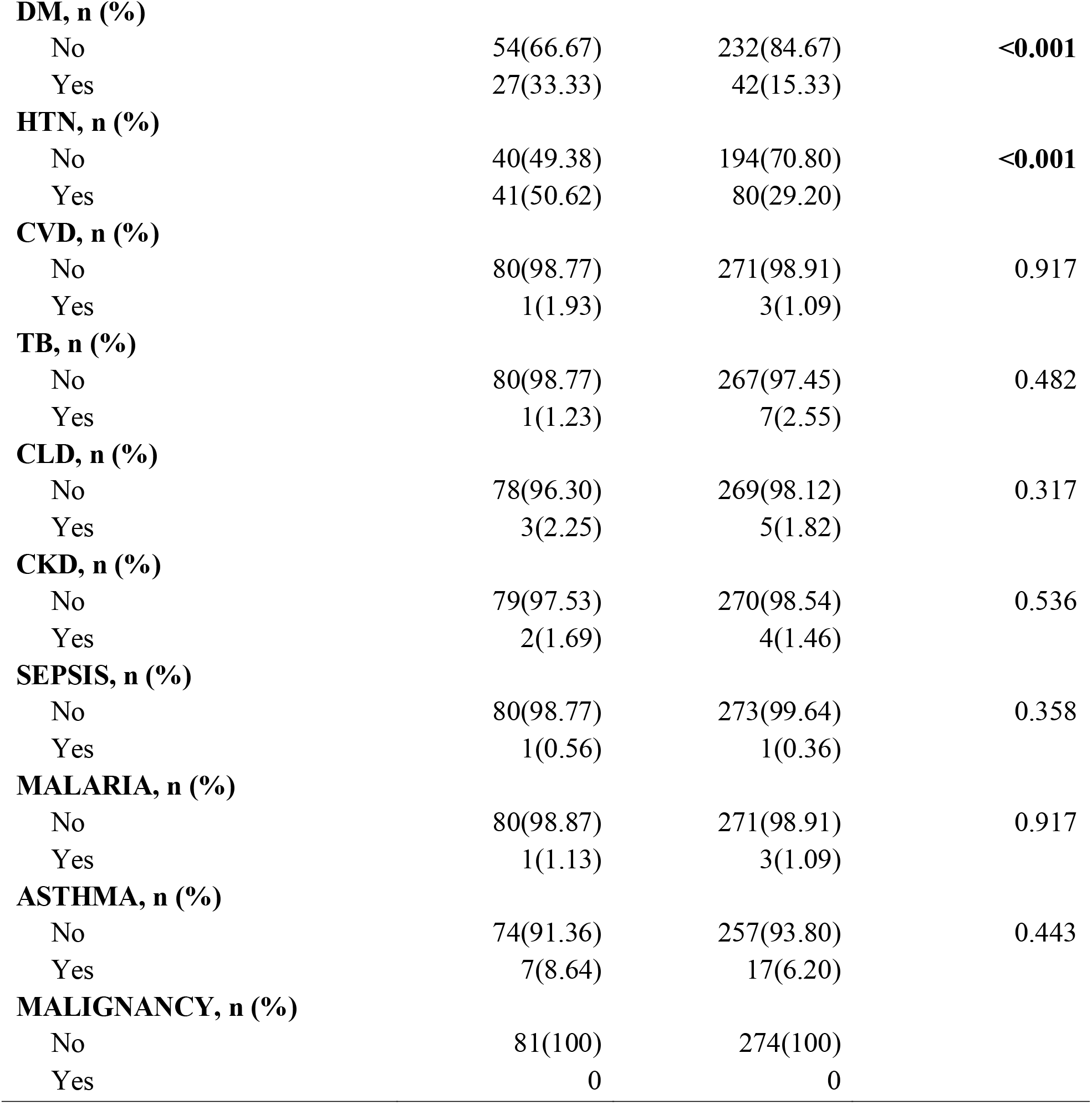
Bivariate analysis of the relationship between the outcome and demographics, clinical characteristics and underlying condition.

### Unadjusted factors associated with COVID-19 outcomes

Table IV shows results of unadjusted logistic regression analysis of factors associated with COVID-19 outcomes. There is significant association between the age of the client with COVID-19 and the outcome, an increase in unit of age was significantly associated with approximately 6% increased chance of COVID-19 outcome death (OR 1.06, 95% CI 1.04-1.08; P <0.001). Clients presenting with shortness of breath on admission was associated with approximately 78% increased chance of COVID-19 outcome death. It was observed that for clients with underlying conditions such as HIV positive status (OR 3.54, 95% CI 1.49-8.41; p= 0.004), Diabetes mellitus (OR 2.76, 95% CI 1.50-4.87; p < 0.001) and Hypertension (OR 2.49, 95% CI 1.50-4.13; p < 0.001) had an increased chance of COVID-19 outcome death.

**Table IV:** Bivariate Unadjusted analysis of factors associated with COVID-19 outcomes

| Unadjusted analysis |  |  |  |
| --- | --- | --- | --- |
| Characteristics | OR | 95% CI | p-value |
| ***Demographics*** |  |  |  |
| **Age** | 1.06 | 1.04-1.08 | **<0.001** |
| **Gender** |  |  |  |
| Male | ref |  |  |
| Female | 0.59 | 0.34-1.01 | 0.056 |
| ***Clinical characteristics*** |  |  |  |
| **Running nose** |  |  |  |
| No | ref |  |  |
| Yes | 1.3 | 0.23-0.38 | **<0.001** |
| **Shortness of breath** |  |  |  |
| No | ref |  |  |
| Yes | 1.78 | 1.07-2.95 | **0.025** |
| ***Conditions*** |  |  |  |
| **HIV** |  |  |  |
| Negative | ref |  |  |
| Positive | 3.54 | 1.49-8.41 | **0.004** |
| Unknown | 1.61 | 1.49-8.41 | 0.15 |
| **DM** |  |  |  |
| No | ref |  |  |
| Yes | 2.76 | 1.57-4.87 | **<0.001** |
| **HTN** |  |  |  |
| No | ref |  |  |
| Yes | 2.49 | 1.50-4.13 | **<0.001** |

### Adjusted factors associated with COVID-19 Outcomes

In Table V we observed that a one-year increase in age was significantly associated with a 60% increased odds of death as a COVID-19 outcome (OR 1.06; 95% CI 1.04-1.08; P <0.001). Females had a significant 37% reduced chance of COVID-19 outcome death compared with their male counterparts who had an increased odd (OR 0.37; 95% CI 0.18-0.67; p = 0.002). Clients with a positive HIV status had a three times chance of a likelihood to die as compared to those with a negative HIV status (OR 3.04; 95% CI 1.10-8.44; P= 0.032). Individuals who had diabetes mellitus as an underlying condition had two-times odd of dying (OR 2.16, 95% CI 1.05-4.43).

**Table V:** Multivariable analysis of factors associated with COVID-19 outcomes

| Characteristics | Adjusted Analysis |  |  |
| --- | --- | --- | --- |
|  | OR | 95% CI | p-value |
| **Age** | 1.06 | 1.04-1.08 | **<0.001** |
| **Gender** |  |  |  |
| Male | ref |  |  |
| Female | 0.37 | 0.18-0.67 | **0.002** |
| **HIV** |  |  |  |
| Negative | ref |  |  |
| Positive | 3.04 | 1.10-8.44 | **0.032** |
| Unknown | 1.28 | 0.60-2.75 | 0.516 |
| **DM** |  |  |  |
| No | Ref |  |  |
| Yes | 2.16 | 1.05-4.43 | **0.036** |

## Discussion

The findings depict the results of COVID-19 clients admitted at Livingstone Teaching Hospital, a period from January 2020 to September 2021. Routine screening was conducted in which demographics and clinical characteristics were recorded. Age was not normally distributed, the older population were the most affected with an increased chance of mortality [mean (48.5)]. Our findings contradicted the findings of a study done in South Korea that focused on disparities in Age specific morbidity and mortality from SARS-CoV-2 in which they observed a high morbidity and mortality to be among the young population [12].

During the earlier phase of the COVID-19 outbreak, the diagnosis of the coronavirus was complicated by the diversity in symptoms and a number of clinical presentations [1]. Our study captured the following symptoms; Cough 180 (50.7), Fever 138 (38.87%) chest pain 123(34.65%), shortness of breath 121(34.08%), headache 74(20.85%), loss of taste 24(6.67%), loss of smell 27 (7.63%), diarrhea 22 (6.23%), running nose 2 (0.56%), loss of appetite 34 (9.58%). Gastrointestinal symptoms in COVID-19 clients admitted at Livingstone Teaching Hospital COVID-19 ward were uncommon 22(6.23%), a similar pattern in a study done by Guan and another by Jiang in China [1,13]. However, a different pattern was observed by Wang and colleagues in which a higher proportion of cases presented with gastrointestinal symptoms including diarrhea and nausea 14 (10%) was observed (Wang et al., 2020) [14]. The pre-existing and co existing conditions of Diabetes mellitus and hypertension in a client proved to be a high risk factor in COVID-19 outcomes as observed. Similar findings of a previous studies conducted [2,15,16].

Table 31, shows the relationship between COVID-19 outcome and demographics, clinical characteristics as well as underlying conditions. It was observed that age was a significant risk factor of COVID-19 outcome death, the majority of the patients that were admitted and died were older than patients that survived (median age; 65 vs 45; p <0.001). HIV status was a significant risk factor to COVID-19 outcome death. Our findings indicate that the percentage of clients admitted with a known HIV positive status that died were 18.52% was statistically higher than those that survived were 8.23%, p-value 0.013. These were similar conclusions derived in a study that looked at HIV infection and COVID-19 death: showed that people living with HIV had a higher risk of COVID-19 than those living without HIV after adjusting for age and sex [17].

An unadjusted logistic regression in table 4 depicts the bivariate analysis of demographics, clinical characteristics associated with COVID-19 outcomes. Being Female was a potential protection factor from the COVID-19 outcome death as they were likely to survive as compared to the male gender but is not statistically significant (OR 0.59, 95% CI 0.34-1.01; P=0.056). Similar findings in a study done on sex hormones, gender disparity in COVID-19 observed that there is a sex disparity in COVID-19 clinical outcomes, it showed that the females had a lower infection and were less likely to be hospitalized [18]. In addition, the female gender also was observed to present with a better prognosis and a less mortality rate. This trend has been observed in a number of studies. In China, the Chinese Centre for Disease Control and prevention reported a ratio of male to female to be 2.7:1 [19].

Evidence from the current epidemiological data has indicated a bias towards males with regards to higher susceptibility to both COVID-19 infection as well as clinical outcomes [18]. It is cardinal to note that these observation were similar to what was observed during the Severe Acute Respiratory (SARS) and Middle East Respiratory Syndrome (MERS) epidemics [20]. Several factors may come to play to explain these disparity and the protection that comes with being female, this may include the intrinsic differences that exist in the innate and adaptive immune system. These differences have given the female a protective advantage therefore, exploiting such advantages can drive to developing therapeutic strategies to improve clinical outcomes [18].

Our study included determining the association of the variables to the COVID-19 outcome. Observation made on the clinical presentation were as follows Fever (OR 1.11, 95% CI 0.67-1.83; p=0.695), chest pain (OR 1.07, 95% CI 0.64-1.80; p=0.804), loss of appetite (OR 1.47, 95% CI 0.67-3.21; p=0.335), loss of taste (OR 1.43, 95% 0.57-3.58; p=0.443), cough (OR 1, 95% CI 0.61-1.63; p=0.986) and headache (OR 1.11, 95% CI 0.61-2.03; p=0.728) from these statistics generated from our study it indicated that the mentioned variables were a potential risk to COVID-19 outcomes but not a significant one. The odds ratio suggested a potential risk but however, the p value did not indicate a significance towards the outcome. This however, does not mean these characteristics are to be overlooked. However, shortness of breath (OR 1.78, 95% CI 1.09-2.09; p=0.025) was a significant risk for the COVID-19 outcome, clients that presented with a shortness of breath upon admission had a poor prognosis. An association was established between Diabetes mellitus (OR 2.76, 95% CI 1.50-4.87; p < 0.001), Hypertension (OR 2.49, 95% CI 1.50-4.13; p < 0.001) and the outcome. These conditions were associated with a client ending up in a critical stage of COVID-19 as observed from another study that was conducted in China which observed that hypertension and Diabetes mellitus were seen to lead into a patient’s critical state [21].

Individuals who had diabetes mellitus as an underlying condition had two-times odd of dying (OR 2.16, 95% CI 1.05-4.43) as per our findings, it is noted that many studies have reported Diabetes to be associated with severe outcomes in terms of COVID-19 infection and mortality, however, data is conflicting. Our study has indicated a significant association. Similar findings were observed in a meta-analysis that was conducted by Kumar and colleagues to explore the relationship that existed between diabetes mellitus and COVID-19 outcomes, their results found diabetes to be significantly associated with COVID-19 mortality and a severe COVID-19 infection with a pooled ratio of 1.90 (95% CI: 1.37–2.64; p < 0.01) and 2.75 (95% CI: 2.09–3.62; p < 0.01) respectively. In this study it was indicated that Diabetes in patients with COVID-19 had a two fold increase in mortality as well as severity [22].

A study was done in Italy that looked at the risk factors associated with COVID-19, their findings are similar with our observation. Hypertension was the most frequent comorbidity, and patients with hypertension had significantly decreased survival. Despite this, in the multivariable analysis, hypertension was not an independent factor associated with mortality. Conclusions in our study coincided with an earlier study that was done on the first 100 COVID-19 cases in Zambia, in which they determined in the most prevalent underlying conditions were hypertension and human immunodeficiency virus infection whose proportion was both at 35%. This study also indicated that 20% of the patients in this study had presented with underlying condition [23].

## Limitations

Due to the limited medical resources available, majority of the clients admitted during this period were those with obvious symptoms as most were encouraged to self-isolate. Secondly, this data was only collected at a single center hospital thereby limited sample size. As such the study may have included disproportionately more patients with poor outcomes. Laboratory profiling could not be done as majority of client’s laboratory results could not be extracted.

## Conclusion

The main clinical manifestations of COVID-19 are fever, cough, chest pain and shortness of breath. Older age, male gender was associated with greater odds of COVID-19 infection. Hypertension was found to be the most prevalent underlying condition, however after a multivariate analysis it was observed that hypertension could not be independently increase the odds of COVID-19 outcome death. Therefore, people who were HIV positive, diabetics had an increased odd of succumbing to COVID-19. It is therefore imperative that targeted policies should be considered by the policy makers in order to address this increased risk groups as the pandemic response evolves and this may cover for the preparedness of such pandemics in the future. Individuals that present with comorbidities and clinical features associated with a severe outcome of COVID-19 should be monitored closely. Preventive efforts should be put in place especially to target those who are hypertensive, diabetics and on ART.

## Data Availability

All data produced in the present study are available upon reasonable request to the authors

## Ethical considerations

The study did not require access to human subjects as it is a desk-based project. However, all primary research articles included in this study were thoroughly screened to determine if they met the required ethical requirements and principles.

## AUTHOR CONTRIBUTIONS

PS

- Work conception. Data acquisition, analysis and interpretation.
- Manuscript drafting.
- Approval of final manuscript.
- Accountable for all aspects of the work regarding its accuracy or integrity.

MZ

- Manuscript drafting.
- Critical revisions for intellectual content.
- Data interpretation
- Approval of final manuscript.
- Accountable for all aspects of the work regarding its accuracy or integrity.

PJC

- Manuscript drafting.
- Critical revisions for intellectual content.
- Data interpretation
- Approval of final manuscript.
- Accountable for all aspects of the work regarding its accuracy or integrity.

## Acknowledgement

This article is a part of the master thesis submitted to the University of Lusaka in partial fulfilment of the requirements for the degree of Master of Public Health.

